# Investigation of the urinary peptidome to unravel collagen degradation in health and kidney disease

**DOI:** 10.1101/2024.09.30.24314592

**Authors:** Ioanna K Mina, Luis F Iglesias-Martinez, Matthias Ley, Lucas Fillinger, Paul Perco, Justyna Siwy, Harald Mischak, Vera Jankowski

## Abstract

Naturally occurring fragments of collagen type I alpha 1 chain (COL1A1) have been previously associated with chronic kidney disease (CKD), with some fragments showing positive and others negative associations. Using urinary peptidome data from healthy individuals (n=1131) and CKD patients (n=5585) this aspect was investigated in detail. Based on the hypothesis that many collagen peptides are derived not from the full, mature collagen molecule, but from (larger) collagen degradation products, relationships between COL1A1 peptides containing identical sequences were investigated, with the smaller (offspring) peptide being a possible degradation product of the larger (parent) one. The strongest correlations were found for relationships where the parent differed by a maximum of 3 amino acids from the offspring, indicating an exopeptidase-regulated stepwise degradation process. Regression analysis indicated that CKD affects this degradation process. Comparison of matched CKD patients and control individuals (n=612 each) showed that peptides at the start of the degradation process were consistently downregulated in CKD, indicating an attenuation of COL1A1 endopeptidase-mediated degradation. However, as these peptides undergo further degradation, likely mediated by exopeptidases, this downregulation can become less significant or even reverse, leading to an upregulation of later stage fragments and potentially explaining the inconsistencies observed in previous studies.

**Significance Statement:** The current study, after investigating naturally occurring collagen type I alpha 1 chain (COL1A1) degradation fragments in urine, proposes a stepwise degradation process of COL1A1. Initially, the COL1A1 molecule is degraded by endopeptidases, producing larger first fragments, which then undergo further degradation by exopeptidases, resulting in progressively smaller fragments. Notably, the initial COL1A1 fragments are consistently downregulated in chronic kidney disease (CKD), indicating an attenuation of endopeptidase-mediated degradation of COL1A1. This study suggests that the accumulation of collagen in kidney fibrosis results not solely from increased collagen expression, but to a substantial degree from impaired collagen degradation. Additionally, the current study explains inconsistencies in earlier studies associating urinary COL1A1 fragments with fibrotic disease, where mostly negative, but also occasionally positive, associations were observed: While the initial degradation of COL1A1 by endopeptidases is downregulated, subsequent further degradation of these COL1A1-derived peptides by exopeptidases may be increased resulting in some cases in upregulation of smaller peptides. As many of these fragments are valuable biomarkers for fibrosis-related chronic diseases, this study demonstrates the importance of the exact definition of the selected biomarkers, including its C- and N-terminus. Furthermore, understanding the COL1A1 degradation process may provide insights into potential therapeutic targets for treating fibrosis.

## 1. Introduction

Fibrosis is a pathological feature of many chronic diseases, including chronic kidney disease (CKD)[1, 2]. It involves the excessive accumulation of extracellular matrix (ECM) components within tissues, ultimately leading to organ dysfunction[1, 2]. Among these components, collagen type I (COL1) is the most abundant protein in the ECM and a major contributor to fibrotic tissue[2, 3]. COL1 is composed of three chains (two alpha 1 (COL1A1) and one alpha 2 (COL1A2)), forming a characteristic triple helical structure[3]. The biosynthesis of COL1 is a complex process of multiple steps, including the hydroxylation of proline and lysine residues, the triple helix formation, the removal of the N- and C-terminal propeptides, and the assembly of multiple collagen molecules into fibrils stabilised by intermolecular cross-linking[4]. The excessive accumulation of COL1 in fibrotic tissues possibly results from an imbalance between COL1 synthesis and degradation[3, 5].

Degradation fragments of COL1A1 are the most abundant naturally occurring peptides found in urine[6]. Previous research has linked these peptides to CKD, kidney function and fibrosis[7–9]. A landmark study from 2010 investigated for the first time differences in urinary peptides between patients with CKD of different aetiologies and controls, and described the down-regulation of COL1A1 peptides as a prominent component associated with CKD[8]. Mavrogeorgis et al.[9] identified 63 COL1A1 fragments that showed a strong positive correlation (rho > 0.3) with estimated glomerular filtration rate (eGFR), while 6 peptides were negatively correlated (rho < −0.3) with eGFR. Exploring the distribution of urinary peptides in kidney fibrosis, Catanese et al.[7] found 2 COL1A1 peptides increased in patients with fibrotic disease and 3 COL1A1 peptides downregulated. In a previous study, Magalhães et al.[10] reported 3 COL1A1 peptides being significant negatively correlated with fibrosis (rho < −0.3). While these findings generally suggest a reduced degradation of COL1A1 in fibrosis, the inconsistencies observed in the relationship between COL1A1 peptides and kidney disease are obvious and unexplained.

Peptidases are enzymes that degrade proteins by hydrolysing peptide bonds and can be classified based on the reactions they catalyse[11]. Exopeptidases require a free N-terminal amino group, C-terminal carboxyl group, or both, and cleave peptide bonds within three residues of the polypeptide chain terminus[11]. On the other hand, endopeptidases cleave internal peptide bonds within a polypeptide chain, usually quite distant from the N- or C-terminus[11]. Protease activity is crucial for both, the formation of the mature COL1, and its degradation[3]. Specifically, matrix metalloproteinases (MMPs) including MMP-1, MMP-2, MMP-8, MMP-9, MMP-13, MMP-25, and Cathepsin S are involved in the degradation of COL1[3]. The degradation of COL1 by MMPs is regulated at multiple levels, including the transcription and post-translational modifications (PTM) and cellular localisation of the MMPs[3]. Furthermore, MMP activity is also regulated by natural tissue inhibitors of metalloproteinases (TIMPs)[3]. The development of computational tools (such as proteasix) shed some light on how some of these urinary peptides may be generated[12, 13]. Nevertheless, the specific types of proteases involved in generating urinary COL1A1 peptides remain largely unknown.

A previous study demonstrated that many of the identified COL1A1 urinary peptides are derived from the same regions on the COL1A1 molecule[9]. Additionally, numerous larger peptides contain the entire sequences of smaller ones[9]. This observation led to the hypothesis that smaller COL1A1 peptides are not derived directly from the collagen molecule but may originate from the degradation of larger COL1A1 peptides, and that different proteases may be involved in the generation of the larger and smaller peptides, which may be a plausible explanation for the observed apparent inconsistencies in regulation of collagen peptides in fibrotic diseases. Here, we investigated this hypothesis by exploring “parent-offspring peptide relationships,” where the “parent” (the larger peptide) is degraded to produce the “offspring” (the smaller peptide). The resulting data indicate that these peptides may indeed be generated in a stepwise manner, primarily regulated by exopeptidases. Furthermore, by investigating the initial, larger peptides that are potentially directly derived from collagen through endopeptidase activity (“first parents”), we found that these peptides were consistently downregulated in CKD, supporting the hypothesis that kidney fibrosis may result at least in part from attenuated collagen degradation.

## 2. Methods

### 2.1 CE-MS data

The CE-MS analysis is described in detail in previous studies[6, 14, 15]. In short, the analysis was performed using a P/ACE MDQ CE system (Beckman Coulter, Fullerton, CA, USA) coupled with a micro-TOF-MS (Bruker Daltonics, Bremen, Germany). The raw CE-MS data were analysed with MosaFinder software, which assigned signals to a list of 21559 peptides, over 5000 of which have known amino acid sequences[16]. To normalize peptide intensities, 29 collagen fragments, which are generally unaffected by disease, were used as internal standards[17].

### 2.2 Dataset selection

Anonymized urinary peptidome and phenotypic data were retrieved from the Human Urinary Proteome Database[16]. Ethical review and approval were waived for this study (based on the ethics opinion received from the ethics committee of the Hannover Medical School, Germany no. 3116-2016), due to all data being fully anonymized. To study normal degradation pathways, datasets from patients with no evidence of disease and no indication of impaired kidney function (eGFR > 90 mL/min/1.73m^2^, if available) were selected. To investigate changes in CKD, datasets from patients with impaired kidney function (eGFR < 60 mL/min/1.73m^2^) were used (**Supplementary Figure 1**).

For the analysis of peptide regulation in CKD, urinary peptidome datasets from 1000 CKD patients and 1000 non-CKD control individuals were randomly extracted from Human Urinary Proteome Database[16]. These datasets were matched for patients’ age, sex, and blood pressure (**Supplementary Figure 1**).

The analysis focused on the COL1A1 peptides, for which normalised signal intensities were extracted across the selected individuals from the Human Urinary Proteome Database[16]. Intensities of peptides with the same start and stop positions, but with different PTMs (hydroxylation of lysine or proline) were summed up to reflect the total intensity of protein fragments with the same sequence. Only peptides with a frequency > 30% in the healthy cohort (n=1131) were shortlisted for all downstream analyses. The top 10 most abundant peptides with no overlapping sequences in the healthy cohort were selected as “anchor” peptides.

### 2.3 Parent-offspring relationship and statistical analysis

Parent-offspring relationships were determined based on sequence. A relationship was defined if the entire sequence of the offspring peptide was included in the parent peptide sequence.

For each parent-offspring relationship, Spearman’s correlation test was conducted to evaluate the correlation between parent and offspring peptides based on their abundances in the healthy cohort (n=1131). Additionally, a linear model was constructed, with the parent peptide as the independent variable and the offspring peptide as the dependent variable, setting the intercept to zero. This was based on the premise that the absence of the parent peptide would result in the absence of the offspring peptide. The linear models were developed separately for the healthy (n=1131) and CKD (n=5585) cohorts.

To evaluate differences in peptides intensities statistical analysis was performed between matched CKD cases (n=612) and control individuals (n=612) using the non-parametric Mann–Whitney test. Before the analysis missing abundance values were replaced with 0, as applied in previous studies investigating the peptidome regulation[18].

P-values were adjusted using the Benjamini-Hochberg (BH) method[19]. All calculations were completed in R (version 4.3.1) using the cor.test (), lm(), wilcox.test() and p.adjust() functions from the stats package.

### 2.4 Proteases analysis

An in-silico analysis was performed using the peptide-centric tool Proteasix[12] to predict the endopeptidases potentially involved in generating the COL1A1 peptides. Proteasix reconstructed the cleavage sites at the N- and C-terminal sequences of peptides and predicted the endopeptidases involved in their generation based on established protease/cleavage site (CS) associations[20]. The tool offers two modes: one returns predictions based on experimentally observed protease/CS associations, and the other mode returns predicted protease/CS associations[20]. Both observed and predicted modes of the tool were used, considering only results based on human proteases. Results related to digestive enzymes activated only in the acidic conditions of the stomach were omitted, as they were not relevant to this study. The analysis was conducted in 2023, when Proteasix was still operational.

Proteases were assigned as possibly regulating specific parent-offspring peptide relationships based on two criteria: (i) the offspring peptide was predicted to be generated by the protease, and (ii) the parent peptide contained the complete cleavage site recognised by the protease within its sequence.

## 3. Results

### 3.1 Cohort characteristics

The healthy cohort used for investigating degradation pathways consisted of 1131 unique samples. Mean eGFR value in this cohort was 103.7 (SD 10.1) mL/min/1.73 m^2^. The healthy cohort had an average age of 41.4 (SD 14.2) years and included 42% males. The CKD cohort, used to investigate degradation in disease, included 5585 samples, all with an eGFR < 60 (mean eGFR 39.8, SD 13.7) mL/min/1.73 m^2^. Average age was 66.7 (SD 13.3) years, with 64% males in the CKD cohort.

Due to the apparent significant difference in age and sex between the cohorts, and the known associations of age, sex, and blood pressure with the urinary peptidome[21–23], a random sample of 1000 individuals with normal kidney function (eGFR > 90 mL/min/1.73m²) and 1000 individuals with impaired kidney function (eGFR < 60 mL/min/1.73m²) were extracted. These two cohorts were matched for age, blood pressure and sex, resulting in the selection of 612 datasets each. Clinical characteristics of the CKD patients (n=612) and non-CKD control individuals (n=612) used for the peptide regulation analysis are summarized in **Supplementary Table 1**. The study population consisted of 377 male CKD patients and 378 male controls. The mean age was 60.8 (SD 11.1) years for CKD patients and 60.4 (SD 10.3) years for control individuals. Mean diastolic blood pressure was 77.9 (SD 11.1) mmHg for cases and 78.2 (SD 8.5) mmHg for controls, while mean systolic blood pressure was 137.0 (SD 17.2) mmHg and 135.2 (SD 14.3) mmHg for cases and controls, respectively. There were no statistically significant differences in sex, age, or blood pressure between the two groups. Per design, CKD patients had significantly lower eGFR levels as compared to controls (45.5 (SD 12.3) vs 99.8 (SD 8.9) mL/min/1.73 m^2^, p < 0.001).

### 3.2 Parent-offspring peptide relationships indicate an exopeptidase-mediated degradation of COL1A1 peptides

The study design is depicted in **Figure 1**. A total of 788 COL1A1 peptides have been identified in the Human Urinary Proteome Database[16], covering 71% of the full collagen sequence and 95% of the mature collagen sequence. With the exception of two peptides (136-151 and 147-179) that contain N-propeptide sequences, all identified peptides are derived from the mature COL1A1 sequence, consequently reflecting COL1A1 degradation. COL1A1 is highly modified through proline hydroxylation, which introduces a mass change of 16 Da. This PTM occurs during the COL1A1 biosynthesis and appears to have minimal impact on degradation[9], therefore peptides with identical sequences but varying degrees of proline hydroxylation were combined. The 788 COL1Α1 peptides corresponded to 582 unique peptide sequences, with different start and/or stop amino acid position. Of these, 304 peptides were present at frequencies exceeding 30% in the healthy cohort (n=1131 samples). These were considered for the subsequent investigation of COL1A1 degradation processes. Notably, the sequences of many smaller peptides were entirely included in the sequences of larger peptides. This observation suggested a step-wise degradation process where larger peptides may be degraded to smaller ones. Based on the hypothesis that these smaller peptides are generated from larger ones during the COL1A1 degradation, we focused on such “parent - offspring peptide relationships”. A “parent-offspring relationship” is defined here by a larger peptide (the parent) that could be degraded into a smaller peptide (the offspring).

**Figure 1:**
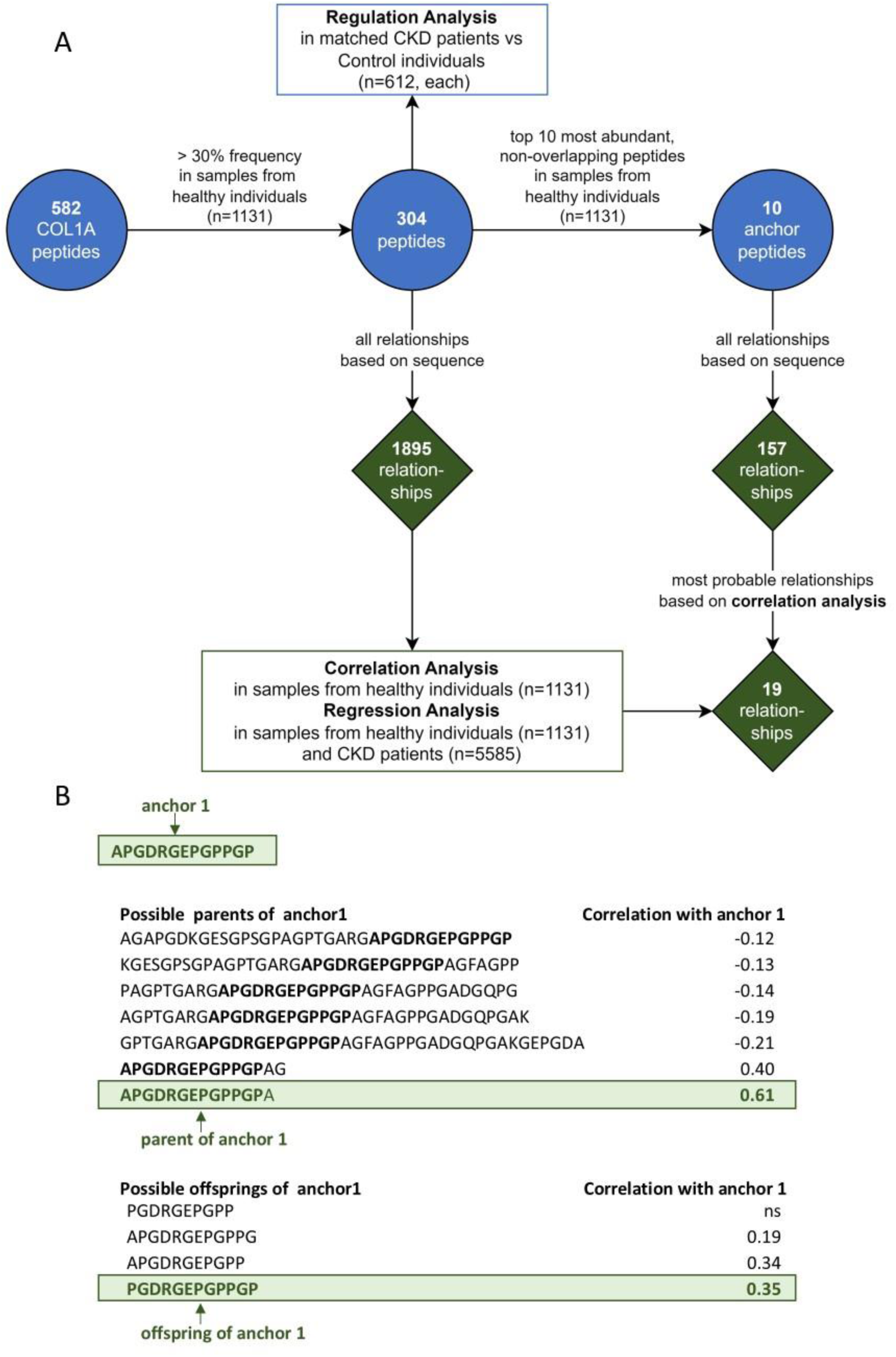
Study Design. (A) Out of the 582 identified COL1A1 urinary peptides, 304 peptides passed a 30% frequency threshold in 1131 samples from healthy individuals, being possibly associated with each other in 1895 parent-offspring relationships based on sequence (where in a relationship the offspring peptide is the degradation product of the parent peptide). For these 304 peptides, the regulation of the peptides in chronic kidney disease (CKD) was investigated using 612 CKD patients and 612 matched control individuals. For the 1895 relationships, the correlations between offspring and parent peptide were investigated in 1131 samples from healthy individuals, and regression analysis between offspring and parent peptides was performed in samples from healthy individuals (n=1131) and CKD patients (n=5585). Because of the large number of peptides and relationships, the top 10 most abundant peptides in samples from healthy individuals (n=1131) with no overlapping sequences were selected and defined as “anchor peptides”, possibly involved in 157 relationships based on sequence. For each anchor peptide, one relationship with the anchor as offspring and one relationship with the anchor as parent were shortlisted based on the highest absolute correlation, leading to 19 relationships (one anchor was not involved in any relationships as a parent). (B) Example of shortlisting the relationships for anchor 1. Out of all the possible parents of anchor 1, the parent peptide that had the highest significant correlation with anchor 1 was selected. The same approach was applied to select the offspring of anchor 1. The selected parent and offspring of anchor 1 also show the closest sequence similarity to the anchor among all the other candidates, with only one amino acid difference, a phenomenon that was also common when investigating the other anchors.

Based on peptide sequences, the 304 peptides were involved in 1895 theoretical parent-offspring peptide relationships, with one peptide being involved in more than one (theoretical) relationships. Investigating this large number of relationships in parallel proved impractical. Therefore, to reduce the high number of peptides and the complexity of their possible relations, the top 10 most abundant COL1A1 peptides in the healthy cohort with non-overlapping sequences were selected for detailed investigation (**Table 1**). Due to their high abundance, these 10 peptides (referred in this manuscript as “anchor”) are assumed to be of higher stability and representative of the degradation process. Since they originate from distinct, non-overlapping regions of the COL1A1 molecule, no parent-offspring relationships can be observed for the anchor peptides themselves. All anchor peptides were potential offsprings of other peptides, and nine of them (excluding anchor 7 (928-946)) may also function as parents. Anchor 6 (819-843) had the highest number of potential parents (25). Anchor 9 (1008-1041) had the lowest number of potential parents (3) but was involved in the most parent-offspring relationships as a parent, potentially generating 21 different peptides.

**Table 1:**
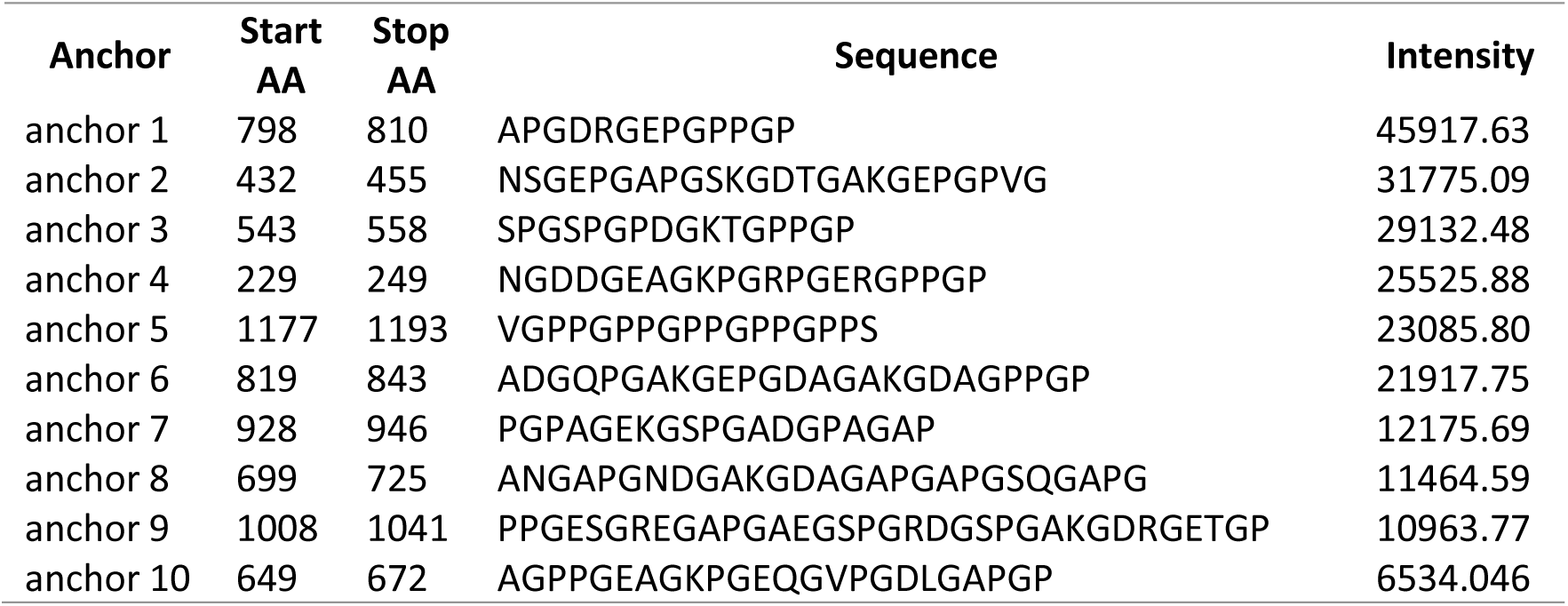
Anchor peptides. Top 10 most abundant non-overlapping collagen type I alpha 1 chain peptides found in at least 30% of healthy samples (n=1131). Peptide intensities were summed for fragments with the same sequence, but different post-translational modifications. Abbreviations: AA-amino acid.

| Anchor | Start<br>AA | Stop<br>AA | Sequence | Intensity |
| --- | --- | --- | --- | --- |
| anchor 1 | 798 | 810 | APGDRGEPGPPGP | 45917.63 |
| anchor 2 | 432 | 455 | NSGEPGAPGSKGDTGAKGEPGPVG | 31775.09 |
| anchor 3 | 543 | 558 | SPGSPGPDGKTGPPGP | 29132.48 |
| anchor 4 | 229 | 249 | NGDDGEAGKPGRPGERGPPGP | 25525.88 |
| anchor 5 | 1177 | 1193 | VGPPGPPGPPGPPGPPS | 23085.80 |
| anchor 6 | 819 | 843 | ADGQPGAKGEPGDAGAKGDAGPPGP | 21917.75 |
| anchor 7 | 928 | 946 | PGPAGEKGSPGADGPAGAP | 12175.69 |
| anchor 8 | 699 | 725 | ANGAPGNDGAKGDAGAPGAPGSQGAPG | 11464.59 |
| anchor 9 | 1008 | 1041 | PPGESGREGAPGAEGSPGRDGSPGAKGDRGETGP | 10963.77 |
| anchor 10 | 649 | 672 | AGPPGEAGKPGEQGVPGDLGAPGP | 6534.046 |

Among the potential parent and offspring peptides identified for each anchor, we aimed to select the peptide that most likely serves as the parent to the anchor (parent of the anchor) and the peptide that is most likely derived from the anchor (offspring of the anchor). The peptides with the highest degree of correlation (of abundance) to the anchor peptide were chosen as parent or offspring, respectively. The correlation was assessed in the healthy cohort (n=1131 samples). For each anchor peptide, the relationship with the highest significant absolute correlation was chosen, both, for the anchor serving as an offspring and for the anchor being the parent (**Figure 1B**). Consequently, a total of 19 relationships were defined (10 parent-anchor and 9 anchor-offspring). In these selected relationships, the correlations between parent and offspring peptides were predominantly significantly positive, with the exception of anchor 7 (928-946), which exhibited a significant negative correlation with its parent peptide. Importantly, 17 of the 19 shortlisted relationships involved parent and offspring peptides that shared either the same starting or the same ending amino acid and differed by up to 3 amino acids at the other end. This pattern indicated that the offspring peptides were likely generated from the parent peptides through exopeptidase activity (**Table 2** and **Supplementary Table 2**).

**Table 2:**
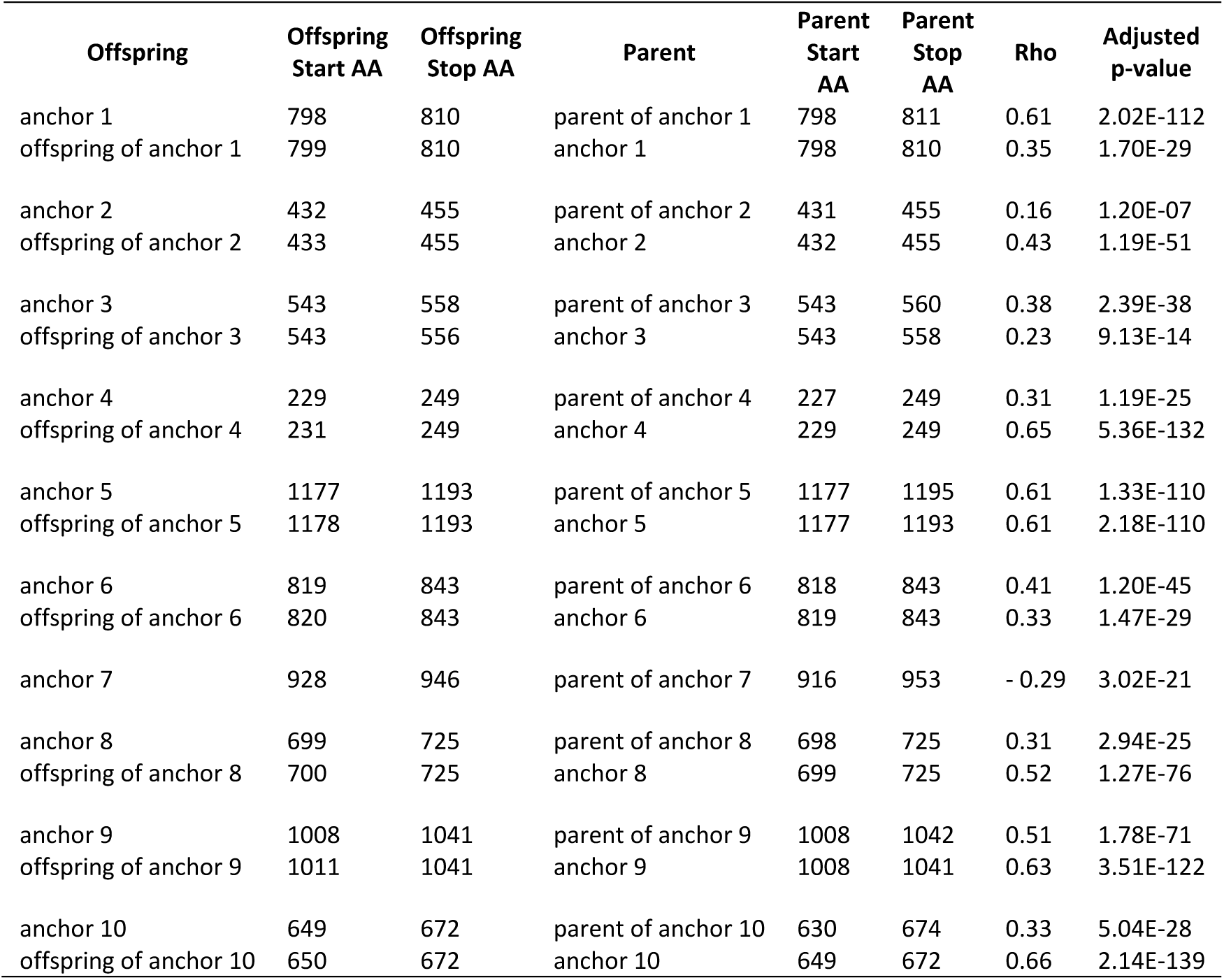
Parent-offspring relationships of the 10 anchor peptides. Each row represents a potential relationship between an offspring peptide and its parent peptide. The most likely parent-offspring relationships involving the anchor peptides, determined by sequence and correlation analysis in 1131 healthy samples, are shown. For each anchor peptide, with the exception of anchor 7 which had no identifiable offspring, one relationship with the anchor as “offspring” and one with the anchor as “parent” were shortlisted. Abbreviations: AA-amino acid.

| Offspring | Offspring Start AA | Offspring Stop AA | Parent | Parent Start AA | Parent Stop AA | Rho | Adjusted p-value |
| --- | --- | --- | --- | --- | --- | --- | --- |
| anchor 1 | 798 | 810 | parent of anchor 1 | 798 | 811 | 0.61 | 2.02E-112 |
| offspring of anchor 1 | 799 | 810 | anchor 1 | 798 | 810 | 0.35 | 1.70E-29 |
| anchor 2 | 432 | 455 | parent of anchor 2 | 431 | 455 | 0.16 | 1.20E-07 |
| offspring of anchor 2 | 433 | 455 | anchor 2 | 432 | 455 | 0.43 | 1.19E-51 |
| anchor 3 | 543 | 558 | parent of anchor 3 | 543 | 560 | 0.38 | 2.39E-38 |
| offspring of anchor 3 | 543 | 556 | anchor 3 | 543 | 558 | 0.23 | 9.13E-14 |
| anchor 4 | 229 | 249 | parent of anchor 4 | 227 | 249 | 0.31 | 1.19E-25 |
| offspring of anchor 4 | 231 | 249 | anchor 4 | 229 | 249 | 0.65 | 5.36E-132 |
| anchor 5 | 1177 | 1193 | parent of anchor 5 | 1177 | 1195 | 0.61 | 1.33E-110 |
| offspring of anchor 5 | 1178 | 1193 | anchor 5 | 1177 | 1193 | 0.61 | 2.18E-110 |
| anchor 6 | 819 | 843 | parent of anchor 6 | 818 | 843 | 0.41 | 1.20E-45 |
| offspring of anchor 6 | 820 | 843 | anchor 6 | 819 | 843 | 0.33 | 1.47E-29 |
| anchor 7 | 928 | 946 | parent of anchor 7 | 916 | 953 | - 0.29 | 3.02E-21 |
| anchor 8 | 699 | 725 | parent of anchor 8 | 698 | 725 | 0.31 | 2.94E-25 |
| offspring of anchor 8 | 700 | 725 | anchor 8 | 699 | 725 | 0.52 | 1.27E-76 |
| anchor 9 | 1008 | 1041 | parent of anchor 9 | 1008 | 1042 | 0.51 | 1.78E-71 |
| offspring of anchor 9 | 1011 | 1041 | anchor 9 | 1008 | 1041 | 0.63 | 3.51E-122 |
| anchor 10 | 649 | 672 | parent of anchor 10 | 630 | 674 | 0.33 | 5.04E-28 |
| offspring of anchor 10 | 650 | 672 | anchor 10 | 649 | 672 | 0.66 | 2.14E-139 |

### 3.3 Possible alterations of exopeptidase-mediated degradation of COL1A1 peptides in CKD

Next, the regulation of anchors and their corresponding parent and offspring peptides, as selected via correlation analysis, was investigated in the context of CKD. For this purpose, peptide abundances were compared between matched cohorts of healthy individuals and patients with CKD, with matching criteria including age, sex, and blood pressure (n=612 each). Of the 10 parent peptides, 8 were significantly downregulated in CKD, while one was significantly upregulated. Of the 10 anchor peptides, 5 were significantly downregulated and one was significantly upregulated. Similarly, among the 9 offspring peptides, 5 were significantly downregulated, and one was significantly upregulated. Overall, a predominance of downregulation in CKD was observed, with this effect being more pronounced at the parent level. In relationships where both, a parent and an offspring peptide, showed significant changes, the regulation trends for the parent and offspring peptides generally aligned. The only exception was the relationship involving anchor 1 (798-810), where the parent peptide was downregulated, while anchor 1 itself was upregulated in CKD (**Table 3** and **Supplementary Table 2**).

**Table 3:**
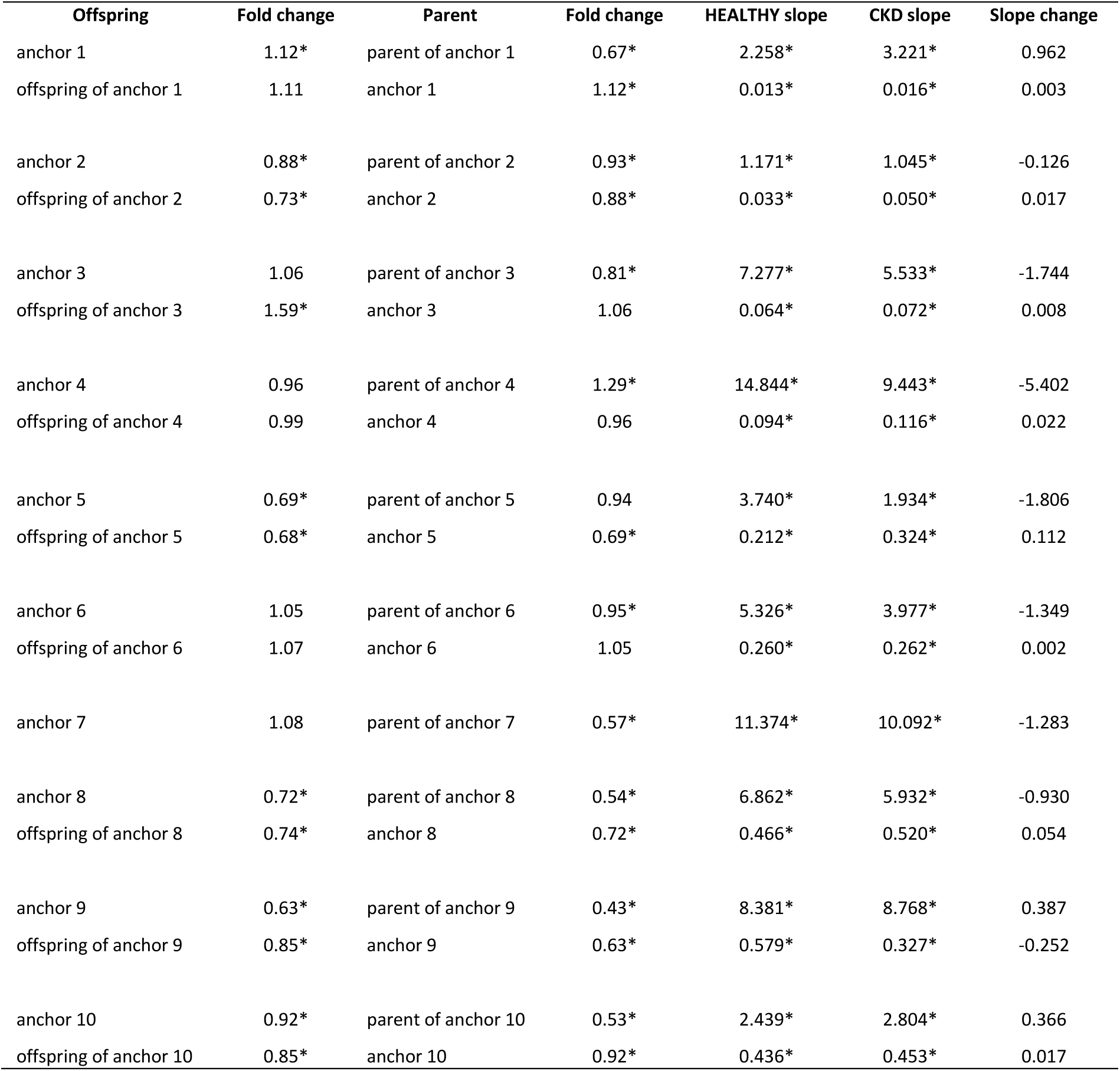
Regulation and regression analysis of anchor parent-offspring relationships in chronic kidney disease (CKD) Each row represents a potential relationship between an offspring peptide and its parent peptide, selected from the most likely relationships involving the anchor peptides. The table shows the regulation of both parent and offspring peptides in CKD (CKD vs Control patients, n=612, each) and regression analysis slope between the parent and offspring peptide in each relationship in healthy (n=1131) and CKD (n=5585) samples. Fold changes are calculated by dividing average value of peptide intensity in cases and average value of peptide intensity in controls. Slope changes are calculated by subtracting healthy slope from CKD slope. Significant fold changes and slopes (p<0.05, Benjamini-Hochberg adjustment) are marked with an asterisk.

| Offspring | Fold change | Parent | Fold change | HEALTHY slope | CKD slope | Slope change |
| --- | --- | --- | --- | --- | --- | --- |
| anchor 1 | 1.12* | parent of anchor 1 | 0.67* | 2.258* | 3.221* | 0.962 |
| offspring of anchor 1 | 1.11 | anchor 1 | 1.12* | 0.013* | 0.016* | 0.003 |
| anchor 2 | 0.88* | parent of anchor 2 | 0.93* | 1.171* | 1.045* | -0.126 |
| offspring of anchor 2 | 0.73* | anchor 2 | 0.88* | 0.033* | 0.050* | 0.017 |
| anchor 3 | 1.06 | parent of anchor 3 | 0.81* | 7.277* | 5.533* | -1.744 |
| offspring of anchor 3 | 1.59* | anchor 3 | 1.06 | 0.064* | 0.072* | 0.008 |
| anchor 4 | 0.96 | parent of anchor 4 | 1.29* | 14.844* | 9.443* | -5.402 |
| offspring of anchor 4 | 0.99 | anchor 4 | 0.96 | 0.094* | 0.116* | 0.022 |
| anchor 5 | 0.69* | parent of anchor 5 | 0.94 | 3.740* | 1.934* | -1.806 |
| offspring of anchor 5 | 0.68* | anchor 5 | 0.69* | 0.212* | 0.324* | 0.112 |
| anchor 6 | 1.05 | parent of anchor 6 | 0.95* | 5.326* | 3.977* | -1.349 |
| offspring of anchor 6 | 1.07 | anchor 6 | 1.05 | 0.260* | 0.262* | 0.002 |
| anchor 7 | 1.08 | parent of anchor 7 | 0.57* | 11.374* | 10.092* | -1.283 |
| anchor 8 | 0.72* | parent of anchor 8 | 0.54* | 6.862* | 5.932* | -0.930 |
| offspring of anchor 8 | 0.74* | anchor 8 | 0.72* | 0.466* | 0.520* | 0.054 |
| anchor 9 | 0.63* | parent of anchor 9 | 0.43* | 8.381* | 8.768* | 0.387 |
| offspring of anchor 9 | 0.85* | anchor 9 | 0.63* | 0.579* | 0.327* | -0.252 |
| anchor 10 | 0.92* | parent of anchor 10 | 0.53* | 2.439* | 2.804* | 0.366 |
| offspring of anchor 10 | 0.85* | anchor 10 | 0.92* | 0.436* | 0.453* | 0.017 |

To investigate whether the observed changes in peptide abundance may be the result of altered degradation processes in CKD, separate regression analyses were performed for the healthy cohort (n=1131) and the CKD cohort (n=5585). Linear models were developed for each parent-offspring peptide pair, with the parent peptide as the independent variable and the offspring peptide as the dependent variable. The models demonstrated good predictive performance, with R^2^ values ranging from 0.37 to 0.88 in the healthy cohort and from 0.19 to 0.78 in CKD. The lower R^2^ values in CKD likely are the result of increased variability in the disease state (**Supplementary Table 2**). All regression slopes in both healthy and CKD were significantly positive, indicating a positive relationship between parent and offspring peptides. Notably, slopes for relationships between anchors and their parents were consistently larger than those between anchors and their offsprings in both conditions. This finding of low degradation levels of the anchor peptides to produce their offspring explains the high abundance and stability of anchor peptides in urine. In CKD, 8 out of 9 slopes representing this low anchor degradation were upregulated, though the difference between healthy and CKD was minimal (≤ 0.12). Conversely, 7 out of 10 slopes characterizing relationships between parent of anchors and anchors decreased in CKD, with reductions ranging from −5.4 to −0.13, indicating a general attenuation of degradation in disease. One of the exceptions was the relationship between anchor 1 and its parent, where the slope increased by 0.96, resulting in the observed downregulation of the parent of anchor 1 and the upregulation of anchor 1 (798-810) in CKD (**Table 3** and **Supplementary Table 2**).

### 3.4 Degradation of COL1A1 is downregulated in CKD, potentially due to altered endopeptidase activity

The observed more pronounced downregulation of parent peptides in CKD, compared to the anchors and their offsprings, suggested a reduction in the endopeptidase-mediated degradation of the full COL1A1 molecule. This initial attenuation of degradation may be followed by subsequent degradation steps mediated mainly by exopeptidases, potentially obscuring the impact of the endopeptidase activity. Consequently, some peptides may lose their negative association with CKD or even exhibit a positive association.

To test this hypothesis, we examined the regulation of the detected “first parent” peptides, the first identified peptides derived directly from the COL1A1 molecule that initiate the degradation pathways leading to the anchor peptides. We expected that these first parent peptides would consistently show downregulation in CKD. Starting from the anchor peptides, the most probable parent of each anchor based on the highest absolute correlation in the healthy cohort (n=1131) was identified, as described in **3.2**. This process was repeated iteratively to identify the most probable parent of each parent peptide until the first, most upstream parent for each anchor was determined. The peptide relationships starting from each first parent and leading to each anchor and their offsprings are listed in **Supplementary Table 3** and the entire degradation pathways are depicted in **Figure 2A**.

**Figure 2:**
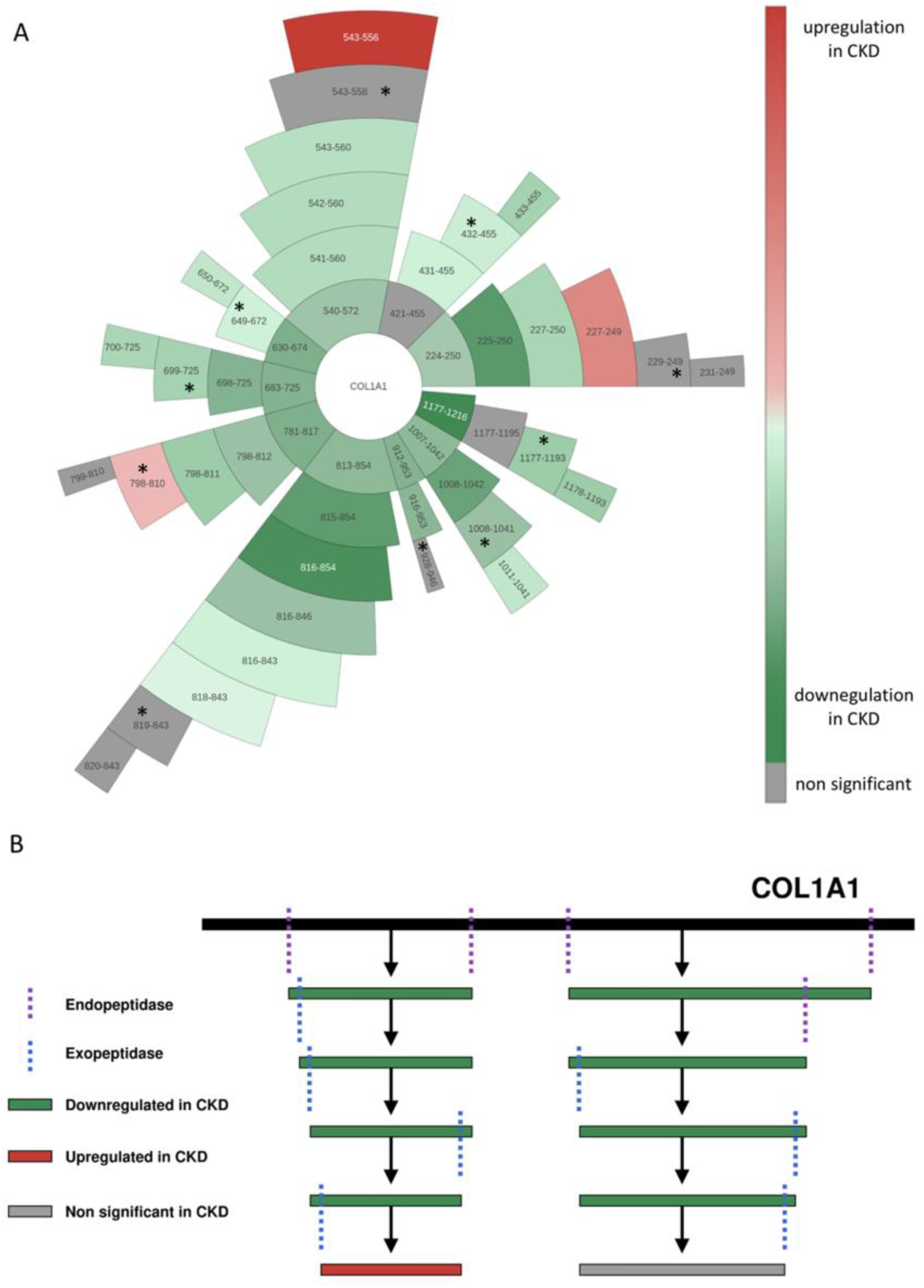
COL1Α1 degradation model. (A) Sunburst plot of COL1A1 parent-offspring relationships using the OntoloViz tool[42] demonstrating the sequential degradation steps of COL1A1 peptides, where one peptide (symbolised by start and stop amino acid) is degraded into another, ultimately leading to the generation of the anchor peptides (marked with *) and their offsprings. These relationships were determined based on sequence and correlation analysis in samples from healthy individuals (n=1131). The regulation of each peptide in chronic kidney disease (CKD) (using CKD patients vs Control individuals, n=612, each) is indicated by colour. The initial layer of peptides after COL1A1 represents the first parent peptides, which were consistently downregulated in CKD. (B) Schematic representation of the COL1A1 degradation process, based on the prediction developed here. Initial degradation of the COL1A1 molecule is likely driven by endopeptidases, producing the generation of the first COL1A1 peptides which are consistently downregulated in CKD. These peptides then undergo multiple degradation steps, potentially initiated by endopeptidases but primarily driven by exopeptidases in the later stages. This stepwise process results in smaller peptides that may lose their statistical significance or even exhibit upregulation in CKD, particularly at the last degradation steps.

On average, each anchor peptide is produced after 3 degradation steps, with anchor 6 (819-843) requiring the most steps (six consecutive parents) and anchor 10 (649-672) the fewest (one parent). In total, 28 relationships were involved in generating all the anchors from their respective first parents. Nineteen of these relationships involved parent and offspring peptides sharing either the same N- or C-terminal amino acid, differing by up to three amino acids at the other end, indicative of exopeptidase activity. The remaining nine relationships, potentially generated by endopeptidases, often involved the first parent peptides, suggesting a more pronounced impact of the endopeptidase activity at the starting steps of the degradation pathways (**Figure 2A** and **Supplementary Table 3**). Proteasix analysis predicted endopeptidases of the MMP and kallikrein families possibly being responsible for five of these nine relationships (**Supplementary Table 3**).

Among the 28 relationships, 25 showed significant positive correlations between parent and offspring peptides, while three, likely generated by endopeptidases based on amino acid differences, showed significant negative correlations. Regression analysis indicated positive slopes for all steps in both healthy and CKD conditions, reflecting the consecutive degradation from parent to offspring peptides. In 6 of the 28 relationships, slopes increased in CKD, while in the remaining 22, slopes decreased, indicating mostly attenuated degradation in CKD (**Supplementary Table 3**).

Of the 38 peptides involved in these 28 relationships, 2 were significantly upregulated, and 30 were significantly downregulated in CKD, showing an overall trend towards downregulation. Importantly, this inconsistency of some peptides being increased in CKD was not observed for the first parents; all nine significantly altered first parents were downregulated in CKD, further supporting the hypothesis of attenuated COL1A1 degradation in CKD (**Figure 2A** and **Supplementary Table 3**).

The first parent peptides either represent fragments of the COL1A1 molecule itself, produced by endopeptidases cleaving within the COL1A1 sequence, or they are fragments of larger COL1A1 peptides not identified by our method. To predict the endopeptidases responsible for generating these first parent peptides, Proteasix analysis was conducted. The analysis indicated that all first parent peptides could potentially be generated by endopeptidase activity, with 5 out of 10 first parents having predicted proteases that cleave both their N- and C-termini. The analysis predominantly predicted members of the MMP as the enzymes responsible for generating these first parent peptides **(Supplementary table 4)**.

In contrast, when the same analysis was applied to the final degradation step in the study (the offsprings of the anchor peptides), endopeptidases were predicted to cleave only 5 out of the 9 offspring peptides. Among these, only the offspring of anchor 3 was predicted to be cleaved at both its N- and C-termini by proteases. These findings further support the hypothesis that endopeptidases are likely involved at the beginning of the COL1A1 degradation process but yet undefined exopeptidases are responsible for the generation of the subsequent, smaller COL1A1 degradation products (**Supplementary table 5**).

## 4. Discussion

Fragments from COL1A1 are the most abundant peptides naturally found in urine[6]. While this observation has not been consistently reported in all manuscripts on this topic, the failure of some reports to correctly identify the collagen peptides almost certainly appears to be due to the authors not taking proline hydroxylation into account, consequently not being able of correctly assigning sequence to the respective peptides[24]. Almost all of the identified COL1A1 peptides are fragments of the mature collagen, likely representing COL1A1 degradation process. COL1A1 peptides have been previously described to be associated with fibrosis-related diseases, including heart failure, CKD, and liver diseases, indicating that the degradation of the COL1 molecule is altered in fibrosis[9, 25, 26]. This association is further supported by the fact that some of these peptides have been directly linked to the level of fibrosis in biopsy[7, 10]. However, the relationship between these peptides and fibrotic diseases is not consistent. Although most of the COL1A1 peptides in these studies are negatively associated with fibrotic disease in the kidney, indicating an attenuated COL1 degradation, this pattern is not consistent, some peptides were also described as being significantly upregulated in disease[7, 9, 10]. Also, an increased abundance of peptides from the COL1A1 termini was found associated with death in the context of heart failure, while peptides from the middle part were decreased[27]. Apparently inconsistent associations of COL1A1 peptides has also been reported in the context of liver fibrosis, with most being highly significantly reduced in fibrosis, however, some being increased[25]. The fact that both decreased, but also increased abundance was observed in disease, in fact even in the same samples and both directional changes being highly significant, indicate that this observation cannot be the result of variability, but must be based on some consistent underlying mechanism. Towards a potential explanation of these observed apparent inconsistencies, the current study investigated possible steps of the COL1A1 degradation process leading to peptide generation and how this degradation process is affected in CKD.

Examining the parent-offspring relationships among COL1A1 peptides, we developed and now propose a model based on sequential degradation processes, where peptides are derived from one another through multiple steps. Correlation analysis between parent and offspring peptides suggested that in most cases each degradation step involves the removal of up to three amino acids from one end of the parent peptide to generate the offspring peptide. This stepwise process continues, with each offspring peptide subsequently becoming a parent peptide itself, undergoing further cleavage of up to three amino acids from one end to produce the next offspring peptide, and continuing in this manner through successive degradation steps. The observed small amino acid differences between parent and offspring peptides in these relationships strongly indicate that these sequential degradation steps may be performed by exopeptidases, enzymes that hydrolyse peptide bonds at the termini of peptide molecules, typically cleaving up to three amino acids at a time[11] (**Figure 2B**). Regression analysis supported these findings, also indicating that while the degradation process from parent to offspring peptide persists in CKD, its rate may be altered. Specifically, a more pronounced downregulation of the exopeptidase-mediated degradation process was observed in CKD.

A previous study from Siwy et al.[28] has demonstrated the role of exopeptidase activity in generating collagen peptides in urine. Specifically, in patients with type II diabetes, inhibiting dipeptidyl peptidase-4 (DPP4), an enzyme that cleaves X-proline dipeptides from the N-terminus of polypeptides, using the DPP4 inhibitor linagliptin, led to changes in numerous urinary peptides. Most of these peptides were collagen fragments and analysis of their sequences indicated that they were likely cleavage products resulting from DPP4 activity[28]. Interestingly, DPP4 is implicated in fibrosis across various organs, including the skin, kidneys, liver, heart, and lungs, and holds promise as a potential therapeutic target for the treatment of fibrosis[29, 30].

Consistent with previous studies, the abundance of the COL1A1 peptides in CKD showed a general trend towards downregulation, though this trend was not always consistent. Downregulation of peptide abundance was more consistent for the largest peptides, suggesting that the closer a degradation step is to the COL1A1 molecule, the more consistent the downregulation in CKD. To further explore this hypothesis, we traced the larger identified peptides, from which the sequential degradation process begins. These “first parent” peptides likely represent fragments of the COL1A1 molecule itself, produced by endopeptidases that cleave within the COL1A1 sequence. Theoretically they could also be fragments of larger COL1A1 peptides not yet identified, however, we found no evidence for such a scenario. In either case, these first parent peptides, being either direct products of COL1A1 degradation, or closer to this initial degradation than all the other identified peptides, more accurately reflect the initial degradation process of the full COL1A1 molecule. The observed consistent downregulation of these first parent peptides in CKD supports the hypothesis that COL1A1 degradation is attenuated in fibrosis, contributing to the accumulation of COL1A1 in fibrotic tissue (**Figure 2B**).

Applying Proteasix to predict endopeptidases potentially responsible for generating these first parent peptides resulted in predicting matrix metalloproteinases (MMPs) as the main enzymes involved. MMPs are well-studied (ECM) proteases, known for their cleavage of COL1A1 chain and their involvement in various types of fibrosis[2, 3]. However, the association between MMPs and fibrosis is complex and varies depending on the specific MMP and type of fibrosis. While it might be expected that MMPs would attenuate fibrosis by degrading collagen, this does not appear to be the case consistently. In some studies, inhibition of MMPs was reported to even promote fibrosis, possibly through pro-inflammatory pathways[2]. Similarly, TIMPs, which regulate MMP activity, show inconsistent effects on fibrosis: depending on the specific TIMP and tissue type, TIMPs can either exacerbate or reduce fibrosis[2]. Previous studies have linked changes in collagen peptides to dysregulated MMP function in kidney disease[31]. In a study by Metzger et al.[32] changes in COL1A1 fragments in urine from patients with kidney allograft rejection suggested the involvement of MMP-8, with biopsy staining confirming increased MMP-8 presence in rejected allografts. Similarly, in a study by Krochmal et al.[13], differences in urinary peptides between patients with diabetic nephropathy and those with diabetes but no nephropathy were investigated, with COL1A1 fragments showing the most pronounced changes, indicating decreased activity of MMP-2 and MMP-9. This prediction was confirmed by zymography experiments in a diabetic nephropathy mouse model[13].

Apart from MMP dysregulation, differences in collagen crosslinking could result in attenuated collagen degradation in fibrosis. Previous studies have shown that both, the levels and the composition of collagen cross-liniking is altered in fibrosis. Specifically, elevated lysyl hydroxylase expression increases hydroxyallysine cross-links at the expense of allysine cross-links, making collagen less susceptible to degradation[33]. Furthermore, LOX enzymes, also involved in collagen crosslinking, are regarded as important contributors to fibrosis development, with multiple LOX inhibitors being currently examined as potential therapeutic options for many types of fibrosis[34]. The accumulation of advanced glycation end products (AGEs) is another possible factor influencing collagen crosslinking in fibrosis. Increased AGE levels result in increased collagen crosslinking and decreased collagen degradation by MMPs[3, 35, 36].

While increased COL1 production in fibrosis is well-documented, the potential role of attenuated collagen degradation has received less attention[37]. However, it appears increasingly plausible that the molecular basis of fibrosis is in fact the impaired degradation. A failure to resolve the excess extracellular matrix tissue, which may be seen as a result of extended “wound healing”, appears as a plausible cause for fibrosis in this context. If this hypothesis is correct, then therapeutic approaches targeting collagen production will obviously not meet success, as such approaches aim towards the incorrect target. Further research along these lines appears urgently needed.

Urinary peptidome analysis offers a unique advantage in understanding this aspect. By precisely identifying and measuring COL1 degradation products in urine, this method provides deep insights into the degradation process and its regulation during disease progression. Notably, urinary peptidome analysis has enabled developing non-invasive biomarkers for diagnosis, prognosis, and prediction of fibrosis-related chronic diseases, including CKD and cardiovascular disease[5, 38–41]. Previous urinary peptidome studies have investigated the association between COL1A1 peptides and fibrosis-related kidney disease[7, 9, 10]. However, these studies reported inconsistencies in peptide regulation, with some peptides showing positive and others negative associations with kidney disease. Our findings provide an explanation for these apparent inconsistencies. Specifically, we observed that attenuated degradation is consistently evident in all peptides involved in the earliest degradation step, likely regulated by endopeptidases. However, as degradation progresses, multiple factors, including exopeptidase activity, likely influence the process. This could lead to a loss of significance in downregulation or even upregulation in later degradation steps, explaining the inconsistent peptide regulation observed in previous studies (**Figure 2B**). This finding has significant implications for proteomic/peptidomic biomarker development. The data demonstrate that the exact definition of the biomarker, based on N- and C-terminus and possibly also present modifications is of the outmost importance. If the detection system relies on just a short stretch of sequence, then in the case presented here, several different peptides will be combined into one signal, even though their regulation is obviously different, potentially resulting in low reproducibility and sometimes puzzling findings. Future studies investigating collagen-derived biomarkers will benefit from the knowledge of the step-wise COL1A1 degradation, to guide choosing biomarkers of the highest relevance and consistency, likely peptides closer to the COL1A1 molecule at the first steps of degradation.

The findings should be interpreted within the context of the study’s limitations. Firstly, the results are based on statistical assumptions and modelling, not including experimental verification of the COL1A1 degradation process. However, such experimental verification appears extremely challenging, considering the complexity of the process and the multiple proteases potentially being involved. Secondly, the analysis focused on 10 anchor peptides selected for their high frequency and abundance, which may not represent the entire spectrum of COL1A1 degradation products. Third, the protease analysis relies on prediction and may not be fully confirmed in a possible future experimental assessment. While this study provides valuable insights into the complex COL1A1 degradation process and offers an explanation for the observed peptide differences in CKD, further research may be needed to confirm and expand upon these findings.

Collectively, investigating the urinary proteome/peptidome enhances our understanding of the COL1A1 degradation process. This could contribute to the development of more effective biomarkers for fibrosis and support identification of potential therapeutic targets to counteract attenuated collagen degradation.

## Data Availability

The datasets used and/or analysed during the current study are available from the corresponding author on reasonable request.

## Abbreviations

CKD: Chronic kidney disease
COL1: Collagen type I
COL1Α1: Collagen type I alpha 1 chain
COL1A2: Collagen type I alpha 2 chain
ECM: Extracellular matrix
eGFR: Estimated glomerular filtration rate
MMP: Matrix metalloproteinases
PTM: Post-translational modifications
TIMP: Tissue inhibitors of metalloproteinases

## Acknowledgements

IKM was supported by a grant from European Union’s Horizon Europe Marie Skłodowska-Curie Actions Doctoral Networks – Industrial Doctorates Programme (HORIZON – MSCA – 2021 – DN-ID, grant number 101072828). VJ was supported by a grant from the ‘Deutsche Forschungsgemeinschaft’ (DFG, German Research Foundation) through the Transregional Collaborative Research Centre (TRR 219; Project-ID 322900939, subproject S-03, INST 948/4S-1); CRU 5011 project number 445703531, Cost-Action CA 21165, IZKF Multiorgan complexity in Friedreich Ataxia and Phase Transition in Disease 1-1, ERA-PerMed (ERA-PERMED2022-202-KidneySign). HM and JS were supported by the German Federal Ministry of Education and Research (BMBF) via funding to the UPTAKE project (01EK2105B) and SIGNAL (01KU2307, under the frame of ERA PerMed) and via COST-Action PERMEDIK CA21165, supported by COST (European Cooperation in Science and Technology). PP, ML, and LF have received funding from the Austrian Research Promotion Agency (FFG) under grant agreement No. 911422 (Delta4Tech). Views and opinions expressed are those of the authors only and do not necessarily reflect those of the funders.

## References

[1] Zhao, M., Wang, L., Wang, M., Zhou, S., Lu, Y., Cui, H., … Yao, Y. (2022). Targeting fibrosis, mechanisms and cilinical trials. Signal Transduct Target Ther, 7, 206.

[2] Zhao, X., Kwan, J. Y. Y., Yip, K., Liu, P. P., & Liu, F. F. (2020). Targeting metabolic dysregulation for fibrosis therapy. Nat Rev Drug Discov, 19, 57–75.

[3] Devos, H., Zoidakis, J., Roubelakis, M. G., Latosinska, A., & Vlahou, A. (2023). Reviewing the Regulators of COL1A1. Int J Mol Sci, 24.

[4] Yamauchi, M., & Sricholpech, M. (2012). Lysine post-translational modifications of collagen. Essays Biochem, 52, 113–133.

[5] Latosinska, A., Siwy, J., Faguer, S., Beige, J., Mischak, H., & Schanstra, J. P. (2021). Value of Urine Peptides in Assessing Kidney and Cardiovascular Disease. Proteomics Clin Appl, 15, e2000027.

[6] Mavrogeorgis, E., Mischak, H., Latosinska, A., Siwy, J., Jankowski, V., & Jankowski, J. (2021). Reproducibility Evaluation of Urinary Peptide Detection Using CE-MS. Molecules, 26.

[7] Catanese, L., Siwy, J., Mavrogeorgis, E., Amann, K., Mischak, H., Beige, J., & Rupprecht, H. (2021). A Novel Urinary Proteomics Classifier for Non-Invasive Evaluation of Interstitial Fibrosis and Tubular Atrophy in Chronic Kidney Disease. Proteomes, 9.

[8] Good, D. M., Zurbig, P., Argiles, A., Bauer, H. W., Behrens, G., Coon, J. J., … Schmitt-Kopplin, P. (2010). Naturally occurring human urinary peptides for use in diagnosis of chronic kidney disease. Mol Cell Proteomics, 9, 2424–2437.

[9] Mavrogeorgis, E., Mischak, H., Latosinska, A., Vlahou, A., Schanstra, J. P., Siwy, J., … Jankowski, J. (2021). Collagen-Derived Peptides in CKD: A Link to Fibrosis. Toxins (Basel), 14.

[10] Magalhaes, P., Pejchinovski, M., Markoska, K., Banasik, M., Klinger, M., Svec-Billa, D., … Schanstra, J. P. (2017). Association of kidney fibrosis with urinary peptides: a path towards non-invasive liquid biopsies? Sci Rep, 7, 16915.

[11] Rawlings, N. D., Waller, M., Barrett, A. J., & Bateman, A. (2014). MEROPS: the database of proteolytic enzymes, their substrates and inhibitors. Nucleic Acids Res, 42, D503–509.

[12] Klein, J., Eales, J., Zurbig, P., Vlahou, A., Mischak, H., & Stevens, R. (2013). Proteasix: a tool for automated and large-scale prediction of proteases involved in naturally occurring peptide generation. Proteomics, 13, 1077–1082.

[13] Krochmal, M., Kontostathi, G., Magalhaes, P., Makridakis, M., Klein, J., Husi, H., … Vlahou, A. (2017). Urinary peptidomics analysis reveals proteases involved in diabetic nephropathy. Sci Rep, 7, 15160.

[14] Mischak, H., Kolch, W., Aivaliotis, M., Bouyssie, D., Court, M., Dihazi, H., … Vlahou, A. (2010). Comprehensive human urine standards for comparability and standardization in clinical proteome analysis. Proteomics Clin Appl, 4, 464–478.

[15] Mischak, H., Vlahou, A., & Ioannidis, J. P. (2013). Technical aspects and inter-laboratory variability in native peptide profiling: the CE-MS experience. Clin Biochem, 46, 432–443.

[16] Latosinska, A., Siwy, J., Mischak, H., & Frantzi, M. (2019). Peptidomics and proteomics based on CE-MS as a robust tool in clinical application: The past, the present, and the future. Electrophoresis, 40, 2294–2308.

[17] Jantos-Siwy, J., Schiffer, E., Brand, K., Schumann, G., Rossing, K., Delles, C., … Metzger, J. (2009). Quantitative urinary proteome analysis for biomarker evaluation in chronic kidney disease. J Proteome Res, 8, 268–281.

[18] An, D. W., Yu, Y. L., Martens, D. S., Latosinska, A., Zhang, Z. Y., Mischak, H., … Staessen, J. A. (2023). Statistical approaches applicable in managing OMICS data: Urinary proteomics as exemplary case. Mass Spectrom Rev.

[19] Benjamini, Y., & Hochberg, Y. (1995). Controlling the False Discovery Rate: A Practical and Powerful Approach to Multiple Testing. Journal of the Royal Statistical Society: Series B (Methodological), 57, 289–300.

[20] Arguello Casteleiro, M., Klein, J., & Stevens, R. (2016). The Proteasix Ontology. J Biomed Semantics, 7, 33.

[21] Martens, D. S., Thijs, L., Latosinska, A., Trenson, S., Siwy, J., Zhang, Z. Y., … investigators, F. (2021). Urinary peptidomic profiles to address age-related disabilities: a prospective population study. Lancet Healthy Longev, 2, e690–e703.

[22] Mavrogeorgis, E., Kondyli, M., Mischak, H., Vlahou, A., Siwy, J., Rossing, P., … Persu, A. (2024). Multiple urinary peptides are associated with hypertension: a link to molecular pathophysiology. J Hypertens, 42, 1331–1339.

[23] Mina, I. K., Mavrogeorgis, E., Siwy, J., Stojanov, R., Mischak, H., Latosinska, A., & Jankowski, V. (2024). Multiple urinary peptides display distinct sex-specific distribution. Proteomics, 24, e2300227.

[24] Elguoshy, A., Yamamoto, K., Hirao, Y., Uchimoto, T., Yanagita, K., & Yamamoto, T. (2024). Investigating and Annotating the Human Peptidome Profile from Urine under Normal Physiological Conditions. Proteomes, 12.

[25] Bannaga, A. S., Metzger, J., Kyrou, I., Voigtlander, T., Book, T., Melgarejo, J., … Arasaradnam, R. P. (2020). Discovery, validation and sequencing of urinary peptides for diagnosis of liver fibrosis-A multicentre study. EBioMedicine, 62, 103083.

[26] He, T., Mischak, M., Clark, A. L., Campbell, R. T., Delles, C., Diez, J., … Latosinska, A. (2021). Urinary peptides in heart failure: a link to molecular pathophysiology. Eur J Heart Fail, 23, 1875–1887.

[27] He, T., Melgarejo, J. D., Clark, A. L., Yu, Y. L., Thijs, L., Diez, J., … Jankowski, V. (2021). Serum and urinary biomarkers of collagen type-I turnover predict prognosis in patients with heart failure. Clin Transl Med, 11, e267.

[28] Siwy, J., Klein, T., Rosler, M., & von Eynatten, M. (2019). Urinary Proteomics as a Tool to Identify Kidney Responders to Dipeptidyl Peptidase-4 Inhibition: A Hypothesis-Generating Analysis from the MARLINA-T2D Trial. Proteomics Clin Appl, 13, e1800144.

[29] Hu, M. S., & Longaker, M. T. (2016). Dipeptidyl Peptidase-4, Wound Healing, Scarring, and Fibrosis. Plast Reconstr Surg, 138, 1026–1031.

[30] Ohm, B., Moneke, I., & Jungraithmayr, W. (2023). Targeting cluster of differentiation 26 / dipeptidyl peptidase 4 (CD26/DPP4) in organ fibrosis. Br J Pharmacol, 180, 2846–2861.

[31] Klein, J., Bascands, J. L., Mischak, H., & Schanstra, J. P. (2016). The role of urinary peptidomics in kidney disease research. Kidney Int, 89, 539–545.

[32] Metzger, J., Chatzikyrkou, C., Broecker, V., Schiffer, E., Jaensch, L., Iphoefer, A., … Gwinner, W. (2011). Diagnosis of subclinical and clinical acute T-cell-mediated rejection in renal transplant patients by urinary proteome analysis. Proteomics Clin Appl, 5, 322–333.

[33] Piersma, B., & Bank, R. A. (2019). Collagen cross-linking mediated by lysyl hydroxylase 2: an enzymatic battlefield to combat fibrosis. Essays Biochem, 63, 377–387.

[34] Rodriguez-Pascual, F., & Rosell-Garcia, T. (2022). The challenge of determining lysyl oxidase activity: Old methods and novel approaches. Anal Biochem, 639, 114508.

[35] DeGroot, J., Verzijl, N., Budde, M., Bijlsma, J. W., Lafeber, F. P., & TeKoppele, J. M. (2001). Accumulation of advanced glycation end products decreases collagen turnover by bovine chondrocytes. Exp Cell Res, 266, 303–310.

[36] Panwar, P., Butler, G. S., Jamroz, A., Azizi, P., Overall, C. M., & Bromme, D. (2018). Aging-associated modifications of collagen affect its degradation by matrix metalloproteinases. Matrix Biol, 65, 30–44.

[37] Ricard-Blum, S., Baffet, G., & Theret, N. (2018). Molecular and tissue alterations of collagens in fibrosis. Matrix Biol, 68-69, 122–149.

[38] Argiles, A., Siwy, J., Duranton, F., Gayrard, N., Dakna, M., Lundin, U., … Mischak, H. (2013). CKD273, a new proteomics classifier assessing CKD and its prognosis. PLoS One, 8, e62837.

[39] Rossing, K., Mischak, H., Dakna, M., Zurbig, P., Novak, J., Julian, B. A., … Network, P. (2008). Urinary proteomics in diabetes and CKD. J Am Soc Nephrol, 19, 1283–1290.

[40] Tofte, N., Lindhardt, M., Adamova, K., Bakker, S. J. L., Beige, J., Beulens, J. W. J., … investigators, P. (2020). Early detection of diabetic kidney disease by urinary proteomics and subsequent intervention with spironolactone to delay progression (PRIORITY): a prospective observational study and embedded randomised placebo-controlled trial. Lancet Diabetes Endocrinol, 8, 301–312.

[41] Zhang, Z. Y., Thijs, L., Petit, T., Gu, Y. M., Jacobs, L., Yang, W. Y., … Staessen, J. A. (2015). Urinary Proteome and Systolic Blood Pressure as Predictors of 5-Year Cardiovascular and Cardiac Outcomes in a General Population. Hypertension, 66, 52–60.

[42] Ley, M., Heinzel, A., Fillinger, L., Kratochwill, K., & Perco, P. (2023). OntoloViz: a GUI for interactive visualization of ranked disease or drug lists using the MeSH and ATC ontologies. Bioinform Adv, 3, vbad113.

