## Supplementary Figure 1 for "Investigation of the urinary peptidome to unravel collagen degradation in health and kidney disease"

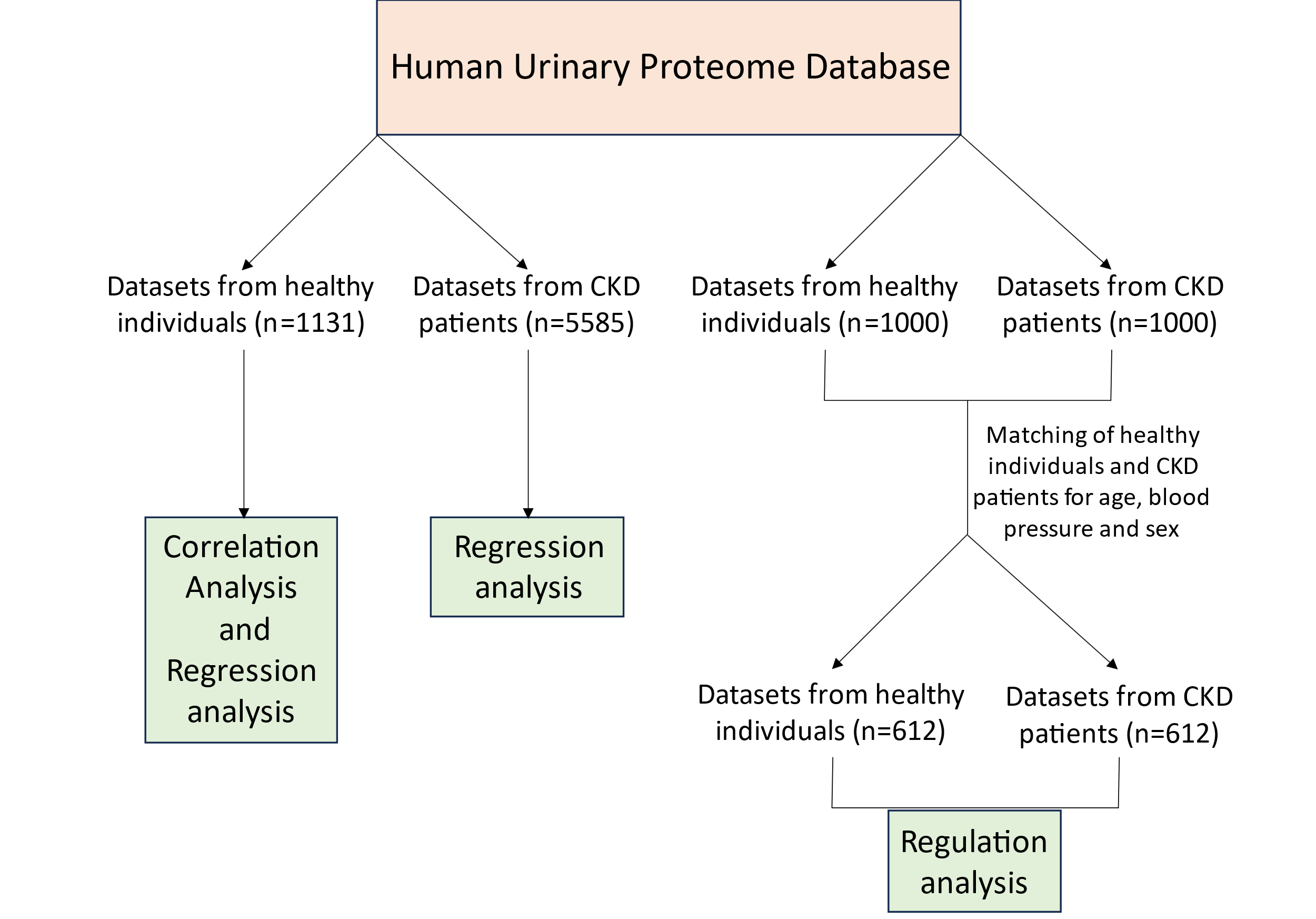

**Supplementary Figure 1: Cohort Selection.** For the investigation of the normal COL1A1 degradation pathway, 1131 datasets from healthy individuals were extracted from the Human Urinary Proteome Database and used for correlation and regression analyses. To examine changes in this process in CKD, 5585 datasets from patients with eGFR<60 ml/min/1.73m² were extracted from the same Database and used for regression analysis. Additionally, to assess changes in peptide abundance between CKD patients and healthy individuals (regulation analysis), 1000 healthy individuals and 1000 CKD patients were randomly extracted from the Human Urinary Proteome Database and matched for age, blood pressure, and sex to eliminate impact by these known confounders [21-23].
